# Effectiveness of the neutralizing antibody sotrovimab among high-risk patients with mild to moderate SARS-CoV-2 in Qatar

**DOI:** 10.1101/2022.04.21.22274060

**Authors:** Ahmed Zaqout, Muna A. Almaslamani, Hiam Chemaitelly, Samar A. Hashim, Ajithkumar Ittaman, Abeir Alimam, Fatma Rustom, Joanne Daghfal, Mohammed Abukhattab, Sawsan AlMukdad, Anvar Hassan Kaleeckal, Ali Nizar Latif, Adeel A. Butt, Roberto Bertollini, Abdullatif Al-Khal, Ali S. Omrani, Laith J. Abu-Raddad

## Abstract

Effectiveness of sotrovimab against severe, critical, or fatal COVID-19 was investigated in Qatar using a case-control study design at a time when BA.2 Omicron subvariant dominated incidence. Adjusted odds ratio of progression to severe, critical, or fatal COVID-19, comparing those sotrovimab-treated to those untreated, was 2.67-fold higher (95% CI: 0.60-11.91).

---

Several monoclonal antibodies (mAbs) against severe acute respiratory syndrome coronavirus 2 (SARS-CoV-2) have been developed for treatment of coronavirus disease 2019 (COVID-19) [1]. One of these is sotrovimab, which significantly reduced the risk of COVID-19 hospitalization and death due to infection with pre-Omicron SARS-CoV-2 variants in a randomized clinical trial [2]. The U.S. Food and Drug Administration (FDA) issued an emergency authorization to permit use of sotrovimab for treatment of mild-to-moderate COVID-19 in patients at high risk of progression to severe COVID-19 [3]. Following FDA guidelines, Qatar’s Ministry of Public Health authorized use of sotrovimab on October 20, 2021. Nonetheless, efficacy of sotrovimab against Omicron (B.1.1.529) subvariants is in question [3]. We estimated real-world effectiveness of sotrovimab against severe, critical, or fatal COVID-19 in Qatar at a time in which most incidence occurred due to the BA.2 Omicron subvariant.

## Methods

### Study population, data sources, and study design

Effectiveness of sotrovimab was investigated using a matched case-control study design among the resident population of Qatar. The study population included patients 12 years of age and older, weighing at least 40 kg, who tested positive for SARS-CoV-2 using real time reverse transcription (RT-qPCR) testing or rapid antigen testing (RAT) between October 20, 2021 and February 28, 2022, and who had at least one risk factor that increases their risk of severe COVID-19 progression per FDA guidelines [3]. No record of COVID-19 vaccination was also considered a risk factor for severe COVID-19 progression.

In compliance with Qatar’s COVID-19 Home Isolation Service guidelines, all outpatient COVID-19 cases were screened for sotrovimab eligibility, and if eligible, were contacted by phone to offer sotrovimab. Clinical and disease outcome data were retrospectively collated for all individuals meeting study eligibility criteria. Notably, Qatar has young, diverse demographics, of that only 9% of its residents are ≥50 years of age, and 89% are expatriates from over 150 countries [4].

Cases (treatment group) included patients who received 500-mg, over 30 minutes infusion of sotrovimab, within 7 days of their PCR-positive or RAT-positive test. Controls were patients who were offered the treatment, but opted not to receive it. Patients were excluded from the treatment group if they showed symptoms of severe COVID-19 (oxygen saturation level <90% or required oxygen supplements) before receiving sotrovimab. Patients were excluded from the control group if they showed signs or symptoms of severe COVID-19 within 7 days of diagnosis.

Cases and controls were exact-matched in a 1:2 ratio by COVID-19 vaccination status, prior infection status, sex, age group, nationality, comorbidity count, and epidemic phase (Delta-dominated incidence versus Omicron-dominated incidence). Vaccination status and prior infection status of cases and controls were ascertained at the time of the PCR or RAT positive test. Prior infection was defined as a positive PCR or RAT test ≥90 days before the PCR or RAT positive test under study [5, 6]. The primary outcome of this study was progression to severe, critical, or fatal COVID-19 among those treated with sotrovimab compared with untreated patients.

The large Omicron-wave exponential-growth phase in Qatar started on December 19, 2021 and peaked in mid-January, 2022 [7, 8]. Accordingly, infections diagnosed prior to December 19, 2021 were classified under the Delta-dominated incidence phase. Infections diagnosed on December 19, 2021 or thereafter were classified under the Omicron-dominated incidence phase. During the Omicron wave, >70% of incident cases were BA.2 infections [7, 8]. The remaining cases were mostly BA.1 cases, with only marginal Delta incidence.

Classification of case severity (acute-care hospitalizations) [9], criticality (intensive-care-unit hospitalizations) [9], and fatality [10] followed World Health Organization guidelines, and assessments were made by trained medical personnel independent of study investigators and using individual chart reviews, as part of a national protocol applied to every hospitalized COVID-19 patient. Details of COVID-19 severity, criticality, and fatality classification are found in Supplementary Text S1.

Every hospitalized COVID-19 patient underwent an infection severity assessment every three days until discharge or death. Individuals who progressed to severe, critical, or fatal COVID-19 were classified based on their worst disease outcome, starting with death [10], followed by critical disease [9], and then severe disease [9]. The study database was linked to the national COVID-19 severity, criticality, and fatality database to ascertain disease outcome for every individual included in this study.

### Statistical analysis

Characteristics of treatment and control groups were described using frequency distributions and measures of central tendency. Group comparisons were performed using standardized mean differences (SMDs), with an SMD of <0.1 indicating adequate matching [11].

Adjusted odds ratios (AORs), comparing odds of progression to severe, critical, or fatal COVID-19 in the treatment versus control group, and associated 95% confidence intervals (CIs) were derived using conditional logistic regression, factoring the matching in the study design.

Analysis was repeated only for a subgroup of patients at higher risk of severe forms of COVID-19. The latter included only immunocompromised individuals (solid organ or hematopoietic stem cell transplant recipients, patients receiving chemotherapy or immunosuppressive treatments, patients with severe immunodeficiency, and HIV patients), unvaccinated individuals, those ≥75 years of age, and pregnant women.

An additional analysis was conducted in which associations with severe, critical, or fatal COVID-19 were investigated using multivariable logistic regression to include all individuals eligible for sotrovimab treatment. AORs and associated 95% CIs were derived.

P-values <0.05 were considered statistically significant. Statistical analyses were performed using Stata software, version 17.0, (Stata-Corp., College Station, Texas).

### Oversight

Hamad Medical Corporation and Weill Cornell Medicine-Qatar Institutional Review Boards approved this retrospective study with a waiver of informed consent. The study was reported following the STROBE guidelines (Supplementary Table 1).

## Results

Of 3,364 individuals with documented SARS-CoV-2 infection who met sotrovimab treatment eligibility during the study period, 519 individuals consented and received the treatment. These constituted the treatment group. The remaining 2,845 individuals, who did not receive the treatment, constituted the controls. A total of 345 individuals in the treatment group were exact-matched in a 1:2 ratio to 583 individuals in the untreated control group.

Supplementary Table 2 shows the baseline characteristics of study participants before and after matching. The median age was 40 years (interquartile range [IQR], 32-50) in the matched treatment group and 39 years (IQR, 33-46) in the matched control group. Less than 9% of cases were ≥60 years of age. Patients were of diverse nationality backgrounds and were predominantly vaccinated females. Most study participants were infected during the Omicron wave.

Supplementary Table 3 shows baseline characteristics of the subgroup at higher risk of severe forms of COVID-19, before and after matching. Matched study groups were overall well balanced in the different analyses (Supplementary Tables 2 and 3).

The AOR of progression to severe, critical, or fatal COVID-19, comparing those treated to those untreated, was 2.67 (95% CI: 0.60-11.91) (Table 1). In the analysis including only the subgroup of patients at higher risk of severe forms of COVID-19, the AOR was 0.65 (95% CI: 0.17-2.48; Table 1).

**Table 1.** Association of sotrovimab treatment with COVID-19 infection severity in matched treatment and control groups.

| Study group | Severe/Critical/Fatal<sup>†</sup> COVID-19 | Mild/asymptomatic infection | AOR (95% CI) |
| --- | --- | --- | --- |
| **Main analysis** |  |  |  |
| Controls<sup>‡</sup> | 3 | 580 | 1.00 |
| Sotrovimab<sup>‡</sup> | 4 | 341 | 2.67 (0.60-11.91) |
| **Subgroup analysis<sup>§</sup>** |  |  |  |
| Controls<sup>§</sup> | 8 | 525 | 1.00 |
| Sotrovimab<sup>§</sup> | 3 | 292 | 0.65 (0.17-2.48) |

Abbreviations: AOR, adjusted odds ratio; CI, confidence interval; COVID-19, coronavirus disease 2019.
<sup>†</sup>Severity [9], criticality [9], and fatality [10] were defined according to the World Health Organization guidelines.
<sup>‡</sup>Cases and controls were exact-matched one-to-two by vaccination status, prior infection status, sex, age group, nationality group, comorbidity count, and epidemic phase. Matching factors are described in detail in Supplementary Table 2.
<sup>§</sup>Subgroup analysis was performed on a subsample that includes only immunocompromised individuals (solid organ or hematopoietic stem cell transplant recipients, patients receiving chemotherapy or immunosuppressive treatments, patients with severe immunodeficiency, and HIV patients), unvaccinated individuals, those $\geq 75$ years of age, and pregnant women.
<sup>§</sup>Cases and controls were exact-matched 1:2 by vaccination status, prior infection status, sex, age group, nationality group, and epidemic phase. Matching factors are described in detail in Supplementary Table 3.

In the additional analysis using multivariable logistic regression on the full sample, the AOR of progression to severe, critical, or fatal COVID-19, comparing those treated to those untreated, was 1.80 (95% CI: 0.61-5.29) (Supplementary Table 4). In the subgroup analysis restricted to only patients at higher risk of severe forms of COVID-19, the AOR was 1.33 (95% CI: 0.44-4.05) (Supplementary Table 5).

## Discussion

There was no evidence for reduced risk of severe forms of COVID-19 among individuals who received sotrovimab treatment per the FDA guidelines. This was also true for the subgroup of patients at higher risk of severe forms of COVID-19. These findings contrast with the effectiveness observed in a randomized control trial and other studies [2, 12-15], but agree with another study that found no evidence of a protective effect [16]. These findings may be explained by the fact that most infections during the study occurred during the Omicron wave and were predominantly due to BA.2 [7, 8]. Recent evidence demonstrated lower neutralizing activity of sotrovimab against the Omicron variant, and particularly against BA.2 [17, 18]. The FDA has recently blocked use of sotrovimab in regions where BA.2 is dominant [3]. An alternative explanation is that those who consented and received the treatment may have perceived a need for this treatment because of poorer underlying health, thereby biasing effectiveness of the treatment.

This study has limitations. While the sample size was not small, it may not be sufficient to precisely measure a small effect, given that we had to adjust the analysis for other factors that affect severity of COVID-19, including age, vaccination, prior infection, comorbidities, and infection variant status. It is possible that many patients declined sotrovimab treatment due to a perception of lower severity of Omicron infections. With the small proportion of Qatar’s population ≥60 years of age [4], our findings may not be generalized to countries in which elderly citizens represent a larger proportion of the population. As an observational study, investigated cohorts were neither blinded nor randomized, so unmeasured or uncontrolled confounding cannot be excluded.

In conclusion, there was no evidence for a protective effect of sotrovimab in reducing COVID-19 severity in a setting dominated by the BA.2 subvariant.

## Data Availability

The dataset of this study is a property of the Qatar Ministry of Public Health that was provided to the researchers through a restricted-access agreement that prevents sharing the dataset with a third party or publicly. Aggregate data are available within the manuscript and its Supplementary information. A limited dataset including the cases and controls and their associated variables that were used in the analysis can be made available for researchers upon request to the corresponding author of this study.

## Notes

## Acknowledgement

We acknowledge the many dedicated individuals at Hamad Medical Corporation, Communicable Disease Center Home Isolation Team, the Ministry of Public Health, the Primary Health Care Corporation, Qatar Biobank, Sidra Medicine, and Weill Cornell Medicine-Qatar for their diligent efforts and contributions to make this study possible.

## Author contributions

AZ, MAA, and ASO co-designed the study, led the database development, and co-wrote the manuscript. HC co-designed the study, performed the statistical analyses, and co-wrote the first draft of the article. LJA co-designed the study, led the statistical analyses, and co-wrote the first draft of the article. All authors contributed to data collection and acquisition, database development, discussion and interpretation of the results, and to the writing of the manuscript. All authors have read and approved the final manuscript.

## Supplementary Material

**Supplementary Text S1. Coronavirus disease 2019 severity, criticality, and fatality classification**

Severe Coronavirus Disease 2019 (COVID-19) disease was defined per the World health Organization (WHO) classification as a severe acute respiratory syndrome coronavirus 2 (SARS-CoV-2) infected person with “oxygen saturation of <90% on room air, and/or respiratory rate of >30 breaths/minute in adults and children >5 years old (or ≥60 breaths/minute in children <2 months old or ≥50 breaths/minute in children 2-11 months old or ≥40 breaths/minute in children 1–5 years old), and/or signs of severe respiratory distress (accessory muscle use and inability to complete full sentences, and, in children, very severe chest wall indrawing, grunting, central cyanosis, or presence of any other general danger signs)” [1]. Detailed WHO criteria for classifying SARS-CoV-2 infection severity can be found in the WHO technical report [1].

Critical COVID-19 disease was defined per WHO classification as a SARS-CoV-2 infected person with “acute respiratory distress syndrome, sepsis, septic shock, or other conditions that would normally require the provision of life sustaining therapies such as mechanical ventilation (invasive or non-invasive) or vasopressor therapy” [1]. Detailed WHO criteria for classifying SARS-CoV-2 infection criticality can be found in the WHO technical report [1].

COVID-19 death was defined per WHO classification as “a death resulting from a clinically compatible illness, in a probable or confirmed COVID-19 case, unless there is a clear alternative cause of death that cannot be related to COVID-19 disease (e.g. trauma). There should be no period of complete recovery from COVID-19 between illness and death. A death due to COVID-19 may not be attributed to another disease (e.g. cancer) and should be counted independently of preexisting conditions that are suspected of triggering a severe course of COVID-19”. Detailed WHO criteria for classifying COVID-19 death can be found in the WHO technical report [2].

**Supplementary Table 1.**
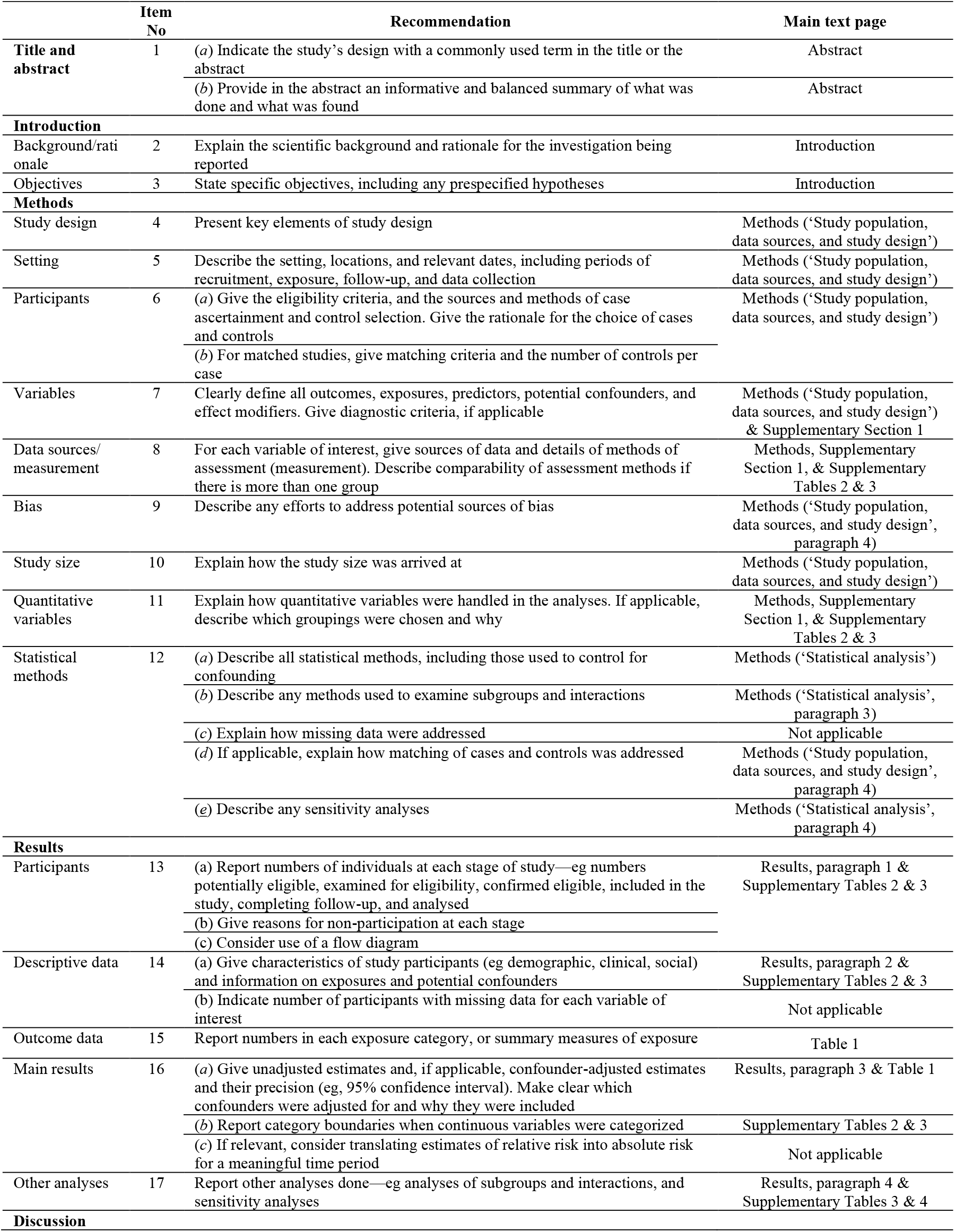
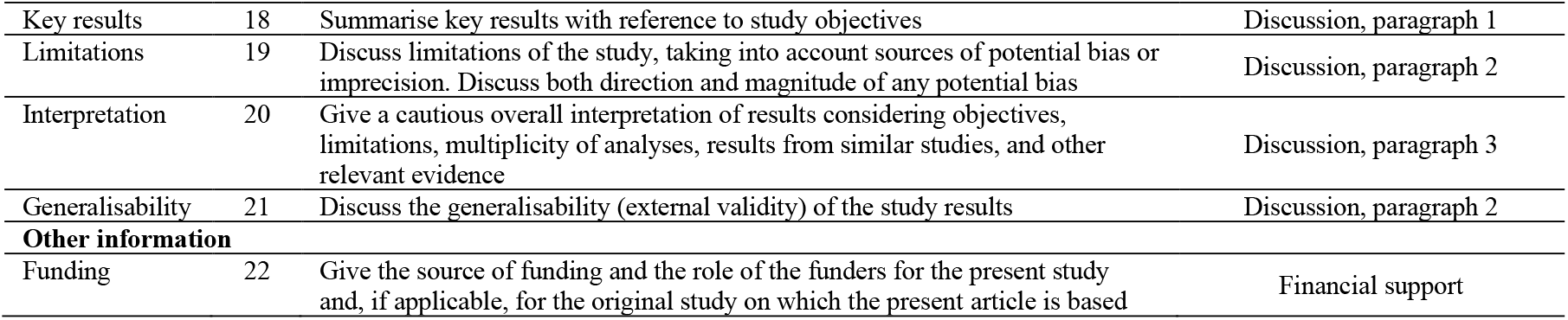
Strengthening the Reporting of Observational Studies in Epidemiology (STROBE) checklist for case-control studies.

|  | Item No | Recommendation | Main text page |
| --- | --- | --- | --- |
| Title and abstract | 1 | (a) Indicate the study’s design with a commonly used term in the title or the abstract | Abstract |
|  |  | (b) Provide in the abstract an informative and balanced summary of what was done and what was found | Abstract |
| Introduction |  |  |  |
| Background/rationale | 2 | Explain the scientific background and rationale for the investigation being reported | Introduction |
| Objectives | 3 | State specific objectives, including any prespecified hypotheses | Introduction |
| Methods |  |  |  |
| Study design | 4 | Present key elements of study design | Methods (‘Study population, data sources, and study design’) |
| Setting | 5 | Describe the setting, locations, and relevant dates, including periods of recruitment, exposure, follow-up, and data collection | Methods (‘Study population, data sources, and study design’) |
| Participants | 6 | (a) Give the eligibility criteria, and the sources and methods of case ascertainment and control selection. Give the rationale for the choice of cases and controls | Methods (‘Study population, data sources, and study design’) |
|  |  | (b) For matched studies, give matching criteria and the number of controls per case |  |
| Variables | 7 | Clearly define all outcomes, exposures, predictors, potential confounders, and effect modifiers. Give diagnostic criteria, if applicable | Methods (‘Study population, data sources, and study design’) & Supplementary Section 1 |
| Data sources/measurement | 8 | For each variable of interest, give sources of data and details of methods of assessment (measurement). Describe comparability of assessment methods if there is more than one group | Methods, Supplementary Section 1, & Supplementary Tables 2 & 3 |
| Bias | 9 | Describe any efforts to address potential sources of bias | Methods (‘Study population, data sources, and study design’, paragraph 4) |
| Study size | 10 | Explain how the study size was arrived at | Methods (‘Study population, data sources, and study design’) |
| Quantitative variables | 11 | Explain how quantitative variables were handled in the analyses. If applicable, describe which groupings were chosen and why | Methods, Supplementary Section 1, & Supplementary Tables 2 & 3 |
| Statistical methods | 12 | (a) Describe all statistical methods, including those used to control for confounding | Methods (‘Statistical analysis’) |
|  |  | (b) Describe any methods used to examine subgroups and interactions | Methods (‘Statistical analysis’, paragraph 3) |
|  |  | (c) Explain how missing data were addressed | Not applicable |
|  |  | (d) If applicable, explain how matching of cases and controls was addressed | Methods (‘Study population, data sources, and study design’, paragraph 4) |
|  |  | (e) Describe any sensitivity analyses | Methods (‘Statistical analysis’, paragraph 4) |
| Results |  |  |  |
| Participants | 13 | (a) Report numbers of individuals at each stage of study—eg numbers potentially eligible, examined for eligibility, confirmed eligible, included in the study, completing follow-up, and analysed | Results, paragraph 1 & Supplementary Tables 2 & 3 |
|  |  | (b) Give reasons for non-participation at each stage |  |
|  |  | (c) Consider use of a flow diagram |  |
| Descriptive data | 14 | (a) Give characteristics of study participants (eg demographic, clinical, social) and information on exposures and potential confounders | Results, paragraph 2 & Supplementary Tables 2 & 3 |
|  |  | (b) Indicate number of participants with missing data for each variable of interest | Not applicable |
| Outcome data | 15 | Report numbers in each exposure category, or summary measures of exposure | Table 1 |
| Main results | 16 | (a) Give unadjusted estimates and, if applicable, confounder-adjusted estimates and their precision (eg, 95% confidence interval). Make clear which confounders were adjusted for and why they were included | Results, paragraph 3 & Table 1 |
|  |  | (b) Report category boundaries when continuous variables were categorized | Supplementary Tables 2 & 3 |
|  |  | (c) If relevant, consider translating estimates of relative risk into absolute risk for a meaningful time period | Not applicable |
| Other analyses | 17 | Report other analyses done—eg analyses of subgroups and interactions, and sensitivity analyses | Results, paragraph 4 & Supplementary Tables 3 & 4 |
| Discussion |  |  |  |
| Key results | 18 | Summarise key results with reference to study objectives | Discussion, paragraph 1 |
| Limitations | 19 | Discuss limitations of the study, taking into account sources of potential bias or imprecision. Discuss both direction and magnitude of any potential bias | Discussion, paragraph 2 |
| Interpretation | 20 | Give a cautious overall interpretation of results considering objectives, limitations, multiplicity of analyses, results from similar studies, and other relevant evidence | Discussion, paragraph 3 |
| Generalisability | 21 | Discuss the generalisability (external validity) of the study results | Discussion, paragraph 2 |
| **Other information** |  |  |  |
| Funding | 22 | Give the source of funding and the role of the funders for the present study and, if applicable, for the original study on which the present article is based | Financial support |

**Supplementary Table 2.** Baseline characteristics of cases and controls.

| Characteristics | Eligible study population | Eligible cases and controls |  |  | Matched cases and controls* |  |  |
| --- | --- | --- | --- | --- | --- | --- | --- |
|  | N=3,364 | Sotrovimab<br>N=519 | Controls<br>N=2,845 | SMD† | Sotrovimab<br>N=345 | Controls<br>N=583 | SMD† |
|  | N (%) | N (%) | N (%) |  | N (%) | N (%) |  |
| **Median age (IQR) — years** | 40 (33-46) | 44 (35-58) | 40 (33-45) | 0.63‡ | 40 (32-50) | 39 (33-46) | 0.20‡ |
| **Age (years)** |  |  |  |  |  |  |  |
| <40 | 1,618 (48.1) | 199 (38.3) | 1,419 (49.9) | 0.64 | 169 (49.0) | 302 (51.8) | 0.07 |
| 40-59 | 1,561 (46.4) | 204 (39.3) | 1,357 (47.7) |  | 147 (42.6) | 241 (41.3) |  |
| ≥60 | 185 (5.5) | 116 (22.4) | 69 (2.4) |  | 29 (8.4) | 40 (6.9) |  |
| **Sex** |  |  |  |  |  |  |  |
| Male | 1,286 (38.2) | 215 (41.4) | 1,071 (37.6) | 0.08 | 124 (35.9) | 195 (33.5) | 0.05 |
| Female | 2,078 (61.8) | 304 (58.6) | 1,774 (62.4) |  | 221 (64.1) | 388 (66.6) |  |
| **Vaccination status§** |  |  |  |  |  |  |  |
| Unvaccinated | 811 (24.1) | 153 (29.5) | 658 (23.1) | 0.19 | 116 (33.6) | 199 (34.1) | 0.04 |
| Two doses | 1,953 (58.1) | 298 (57.4) | 1,655 (58.2) |  | 191 (55.4) | 314 (53.9) |  |
| Three doses | 600 (17.8) | 68 (13.1) | 532 (18.7) |  | 38 (11.0) | 70 (12.0) |  |
| **Prior infection status§** |  |  |  |  |  |  |  |
| No | 3,083 (91.7) | 478 (92.1) | 2,605 (91.6) | 0.02 | 327 (94.8) | 554 (95.0) | 0.01 |
| Yes | 281 (8.4) | 41 (7.9) | 240 (8.4) |  | 18 (5.2) | 29 (5.0) |  |
| **Nationality** |  |  |  |  |  |  |  |
| Qatari | 1,496 (44.5) | 159 (30.6) | 1,337 (47.0) | 0.34 | 103 (29.9) | 186 (31.9) | 0.06 |
| CMW nationalities¶ | 771 (22.9) | 144 (27.8) | 627 (22.0) |  | 83 (24.1) | 127 (21.8) |  |
| Other nationalities | 1,097 (32.6) | 216 (41.6) | 881 (31.0) |  | 159 (46.1) | 270 (46.3) |  |
| **Comorbidity count** |  |  |  |  |  |  |  |
| None | 370 (11.0) | 58 (11.2) | 312 (11.0) | 0.56 | 42 (12.2) | 78 (13.4) | 0.05 |
| 1 | 1,848 (54.9) | 186 (35.8) | 1,662 (58.4) |  | 151 (43.8) | 250 (42.9) |  |
| 2 | 860 (25.6) | 167 (32.2) | 693 (24.4) |  | 105 (30.4) | 181 (31.1) |  |
| ≥3 | 286 (8.5) | 108 (20.8) | 178 (6.3) |  | 47 (13.6) | 74 (12.7) |  |
| **Epidemic phase\*\*** |  |  |  |  |  |  |  |
| Delta-dominated incidence | 460 (13.7) | 198 (38.2) | 262 (9.2) | 0.72 | 112 (32.5) | 152 (26.1) | 0.14 |
| Omicron-dominated incidence | 2,904 (86.3) | 321 (61.9) | 2,583 (90.8) |  | 233 (67.5) | 431 (73.9) |  |

Abbreviations: CMW, craft and manual worker; IQR, interquartile range; SMD, standardized mean difference.
\*Cases (individuals who received the sotrovimab treatment) were exact-matched with up to two controls (individuals who did not receive the sotrovimab treatment) by vaccination status, prior infection status, sex, age group, nationality group, comorbidity count, and epidemic phase. The final sample size thus includes cases that could be matched either to two controls or only to one control. This resulted in small differences between the groups.
†SMD is the difference in the mean of a covariate between groups divided by the pooled standard deviation. An SMD<0.1 indicates optimal balance in matching.
‡SMD is for the mean difference between groups divided by the pooled standard deviation.
§Vaccination status and prior infection status were ascertained at time of infection.
¶These include Bangladeshis, Indians, Nepalese, Pakistanis, Sri Lankans, and Sudanese due to large proportions of these nationals being craft and manual workers.
\*\*Before December 19, 2021 incidence in Qatar was dominated by the Delta variant whereas starting from December 19, 2021 incidence was dominated by the Omicron variant.

**Supplementary Table 3.** Baseline characteristics of the subgroup of patients at higher risk of severe forms of COVID-19*.

| Characteristics | Eligible cases and controls |  |  | Matched cases and controls<sup>†</sup> |  |  |
| --- | --- | --- | --- | --- | --- | --- |
|  | Sotrovimab<br>N=340 | Controls<br>N=1,043 | SMD<sup>‡</sup> | Sotrovimab<br>N=295 | Controls<br>N=533 | SMD<sup>‡</sup> |
|  | N (%) | N (%) |  | N (%) | N (%) |  |
| **Median age (IQR) — years** | 40 (32-56) | 37 (30-44) | 0.54<sup>§</sup> | 39 (32-53) | 36 (30-44) | 0.40<sup>§</sup> |
| **Age (years)** |  |  |  |  |  |  |
| <75 | 308 (90.6) | 1,031 (98.9) | 0.38 | 287 (97.3) | 525 (98.5) | 0.08 |
| ≥75 | 32 (9.4) | 12 (1.2) |  | 8 (2.7) | 8 (1.5) |  |
| **Sex** |  |  |  |  |  |  |
| Male | 100 (29.4) | 274 (26.3) | 0.07 | 71 (24.1) | 118 (22.1) | 0.05 |
| Female | 240 (70.6) | 769 (73.7) |  | 224 (75.9) | 415 (77.9) |  |
| **Condition** |  |  |  |  |  |  |
| Malignancy |  |  |  |  |  |  |
| No | 286 (84.1) | 888 (85.1) | 0.03 | 253 (85.8) | 429 (80.5) | 0.14 |
| Yes | 54 (15.9) | 155 (14.9) |  | 42 (14.2) | 104 (19.5) |  |
| Organ/stem cell transplant |  |  |  |  |  |  |
| No | 304 (89.4) | 1,004 (96.3) | 0.27 | 263 (89.2) | 510 (95.7) | 0.25 |
| Yes | 36 (10.6) | 39 (3.7) |  | 32 (10.9) | 23 (4.3) |  |
| Immunosuppressive treatment |  |  |  |  |  |  |
| No | 256 (75.3) | 969 (92.9) | 0.50 | 225 (76.3) | 493 (92.5) | 0.46 |
| Yes | 84 (24.7) | 74 (7.1) |  | 70 (23.7) | 40 (7.5) |  |
| Severe immunodeficiency |  |  |  |  |  |  |
| No | 338 (99.4) | 1,035 (99.2) | 0.02 | 294 (99.7) | 530 (99.4) | 0.03 |
| Yes | 2 (0.6) | 8 (0.8) |  | 1 (0.3) | 3 (0.6) |  |
| HIV |  |  |  |  |  |  |
| No | 339 (99.7) | 1,042 (99.9) | 0.04 | 294 (99.7) | 532 (99.8) | 0.03 |
| Yes | 1 (0.3) | 1 (0.1) |  | 1 (0.3) | 1 (0.2) |  |
| Pregnancy |  |  |  |  |  |  |
| No | 246 (72.4) | 830 (79.6) | 0.17 | 202 (68.5) | 383 (71.9) | 0.07 |
| Yes | 94 (27.7) | 213 (20.4) |  | 93 (31.5) | 150 (28.1) |  |
| **Vaccination status<sup>¶</sup>** |  |  |  |  |  |  |
| Unvaccinated | 153 (45.0) | 658 (63.1) | 0.37 | 141 (47.8) | 263 (49.3) | 0.04 |
| Two doses | 145 (42.7) | 287 (27.5) |  | 118 (40.0) | 211 (39.6) |  |
| Three doses | 42 (12.4) | 98 (9.4) |  | 36 (12.2) | 59 (11.1) |  |
| **Prior infection status<sup>¶</sup>** |  |  |  |  |  |  |
| No | 312 (91.8) | 953 (91.4) | 0.01 | 276 (93.6) | 502 (94.2) | 0.03 |
| Yes | 28 (8.2) | 90 (8.6) |  | 19 (6.4) | 31 (5.8) |  |
| **Nationality** |  |  |  |  |  |  |
| Qatari | 116 (34.1) | 483 (46.3) | 0.26 | 93 (31.5) | 166 (31.1) | 0.06 |
| CMW nationalities** | 74 (21.8) | 159 (15.2) |  | 62 (21.0) | 101 (19.0) |  |
| Other nationalities | 150 (44.1) | 401 (38.5) |  | 140 (47.5) | 266 (49.9) |  |
| **Epidemic phase<sup>††</sup>** |  |  |  |  |  |  |
| Delta-dominated incidence | 104 (30.6) | 172 (16.5) | 0.34 | 85 (28.8) | 142 (26.6) | 0.05 |
| Omicron-dominated incidence | 236 (69.4) | 871 (83.5) |  | 210 (71.2) | 391 (73.4) |  |

Abbreviations: CMW, craft and manual worker; IQR, interquartile range; SMD, standardized mean difference.
<sup>†</sup>These include immunocompromised individuals (solid organ or hematopoietic stem cell transplant recipients, patients receiving chemotherapy or immunosuppressive treatments, patients with severe immunodeficiency, and HIV patients), unvaccinated individuals, those ≥75 years of age, and pregnant women.
<sup>3</sup>Cases (individuals who received the sotrovimab treatment) were exact-matched with up to two controls (individuals who did not receive the sotrovimab treatment) by vaccination status, prior infection status, sex, age group, nationality group, and epidemic phase. The final sample size thus includes cases that could be matched either to two controls or only to one control. This resulted in small differences between the groups.
<sup>4</sup>SMD is the difference in the mean of a covariate between groups divided by the pooled standard deviation. An SMD<0.1 indicates optimal balance in matching.
<sup>5</sup>SMD is for the mean difference between groups divided by the pooled standard deviation.
<sup>6</sup>Vaccination status and prior infection status were ascertained at time of infection.
<sup>\*\*</sup>These include Bangladeshis, Indians, Nepalese, Pakistanis, Sri Lankans, and Sudanese due to large proportions of these nationals being craft and manual workers.
<sup>††</sup>Before December 19, 2021 incidence in Qatar was dominated by the Delta variant whereas starting from December 19, 2021 incidence was dominated by the Omicron variant.

**Supplementary Table 4.** Associations with COVID-19 infection severity in the full sample including all eligible individuals for sotrovimab treatment.

| Predictors | Sotrovimab |  | Controls |  | Severe/Critical/Fatal* COVID-19 vs. mild/asymptomatic infection |  |  |  |
| --- | --- | --- | --- | --- | --- | --- | --- | --- |
|  | Total sample | Severe/Critical/Fatal* COVID-19 | Total sample | Severe/Critical/Fatal* COVID-19 | Univariable regression analysis |  | Multivariable regression analysis |  |
|  | N (%) | N (%)<sup>†</sup> | N (%) | N (%)<sup>†</sup> | OR (95% CI) | P-value | AOR (95% CI) | P-value |
| **Study group** |  |  |  |  |  |  |  |  |
| Control | -- | -- | 2,845 (100.0) | 10 (0.4) | 1.00 |  | 1.00 |  |
| Sotrovimab | 519 (100.0) | 9 (1.7) | -- | -- | 5.00 (2.02-12.37) | <0.001 | 1.80 (0.61-5.29) | 0.288 |
| **Vaccination status<sup>‡</sup>** |  |  |  |  |  |  |  |  |
| Unvaccinated | 153 (29.5) | 4 (2.6) | 658 (23.1) | 5 (0.8) | 1.00 |  | 1.00 |  |
| Two doses | 298 (57.4) | 4 (1.3) | 1,655 (58.2) | 4 (0.2) | 0.37 (0.14-0.95) | 0.040 | 0.51 (0.17-1.57) | 0.242 |
| Three doses | 68 (13.1) | 1 (1.5) | 532 (18.7) | 1 (0.2) | 0.30 (0.06-1.38) | 0.122 | 0.35 (0.06-1.88) | 0.219 |
| **Prior infection status<sup>‡</sup>** |  |  |  |  |  |  |  |  |
| No | 478 (92.1) | 9 (1.9) | 2,605 (91.6) | 10 (0.4) | 1.00 |  | 1.00 |  |
| Yes | 41 (7.9) | 0 (0.0) | 240 (8.4) | 0 (0.0) | 1.00 (1.00-1.00) | -- | 1.00 (1.00-1.00) | -- |
| **Age (years)** |  |  |  |  |  |  |  |  |
| <40 | 199 (38.3) | 2 (1.0) | 1,419 (49.9) | 5 (0.4) | 1.00 |  | 1.00 |  |
| 40-59 | 204 (39.3) | 2 (1.0) | 1,357 (47.7) | 3 (0.2) | 0.74 (0.23-2.34) | 0.607 | 0.81 (0.24-2.70) | 0.728 |
| ≥60 | 116 (22.4) | 5 (4.3) | 69 (2.4) | 2 (2.9) | 9.05 (3.14-26.10) | <0.001 | 5.68 (1.63-19.77) | 0.006 |
| **Sex** |  |  |  |  |  |  |  |  |
| Male | 215 (41.4) | 4 (1.9) | 1,071 (37.6) | 4 (0.4) | 1.00 |  | 1.00 |  |
| Female | 304 (58.6) | 5 (1.6) | 1,774 (62.4) | 6 (0.3) | 0.85 (0.34-2.12) | 0.728 | 0.75 (0.28-1.98) | 0.558 |
| **Nationality** |  |  |  |  |  |  |  |  |
| Qatari | 159 (30.6) | 3 (1.9) | 1,337 (47.0) | 3 (0.2) | 1.00 |  | 1.00 |  |
| CMW nationalities<sup>§</sup> | 144 (27.7) | 2 (1.4) | 627 (22.0) | 0 (0.0) | 0.65 (0.13-3.21) | 0.593 | 0.70 (0.13-3.72) | 0.678 |
| Other nationalities | 216 (41.6) | 4 (1.9) | 881 (31.0) | 7 (0.8) | 2.52 (0.93-6.82) | 0.070 | 1.91 (0.65-5.60) | 0.240 |
| **Comorbidity count** |  |  |  |  |  |  |  |  |
| None | 58 (11.2) | 0 (0.0) | 312 (11.0) | 4 (1.3) | 1.00 |  | 1.00 |  |
| 1 | 186 (35.8) | 3 (1.6) | 1,662 (58.4) | 5 (0.3) | 0.40 (0.12-1.33) | 0.134 | 1.13 (0.29-4.41) | 0.864 |
| 2 | 167 (32.2) | 1 (0.6) | 693 (24.4) | 1 (0.1) | 0.21 (0.04-1.17) | 0.075 | 0.43 (0.06-2.82) | 0.377 |
| ≥3 | 108 (20.8) | 5 (4.6) | 178 (6.3) | 0 (0.0) | 1.61 (0.43-6.12) | 0.471 | 2.47 (0.44-13.85) | 0.302 |
| **Epidemic phase<sup>¶</sup>** |  |  |  |  |  |  |  |  |
| Delta-dominated incidence | 198 (38.2) | 4 (2.0) | 262 (9.2) | 5 (1.9) | 1.00 |  | 1.00 |  |
| Omicron-dominated incidence | 321 (61.8) | 5 (1.6) | 2,583 (90.8) | 5 (0.2) | 0.17 (0.07-0.43) | <0.001 | 0.35 (0.13-0.96) | 0.041 |

Abbreviations: AOR, adjusted odds ratio; CI, confidence interval; CMW, craft and manual worker; COVID-19, coronavirus disease 2019; OR, odds ratio.
\*Severity [1], criticality [1], and fatality [2] were defined according to the World Health Organization guidelines.
<sup>†</sup>Proportion of those who progressed to severe, critical, or fatal COVID-19 among those in predictor category.
<sup>‡</sup>Vaccination status and prior infection status were ascertained at time of infection.
<sup>§</sup>These include Bangladeshis, Indians, Nepalese, Pakistanis, Sri Lankans, and Sudanese due to large proportions of these nationals being craft and manual workers.
<sup>¶</sup>Before December 19, 2021 incidence in Qatar was dominated by the Delta variant whereas starting from December 19, 2021 incidence was dominated by the Omicron variant.

**Supplementary Table 5.** Associations with COVID-19 infection severity in the subgroup of patients at higher risk of severe forms of COVID-19*.

| Predictors | Sotrovimab |  | Controls |  | Severe/Critical/Fatal<sup>†</sup> COVID-19 vs. mild/asymptomatic infection |  |  |  |
| --- | --- | --- | --- | --- | --- | --- | --- | --- |
|  | Total sample | Severe/Critical/Fatal<sup>†</sup> COVID-19 | Total sample | Severe/Critical/Fatal<sup>†</sup> COVID-19 | Univariable regression analysis |  | Multivariable regression analysis |  |
|  | N (%) | N (%)<sup>‡</sup> | N (%) | N (%)<sup>‡</sup> | OR (95% CI) | P-value | AOR (95% CI) | P-value |
| **Study group** |  |  |  |  |  |  |  |  |
| Control | -- | -- | 1,043 (100.0) | 9 (0.9) | 1.00 |  | 1.00 |  |
| Sotrovimab | 340 (100.0) | 7 (2.1) | -- | -- | 2.42 (0.89-6.53) | 0.083 | 1.33 (0.44-4.05) | 0.618 |
| **Vaccination status<sup>§</sup>** |  |  |  |  |  |  |  |  |
| Unvaccinated | 153 (45.0) | 4 (2.6) | 658 (63.1) | 5 (0.8) | 1.00 |  | 1.00 |  |
| Two doses | 145 (42.6) | 3 (2.1) | 287 (27.5) | 3 (1.0) | 1.26 (0.44-3.55) | 0.668 | 1.52 (0.49-4.70) | 0.468 |
| Three doses | 42 (12.4) | 0 (0.0) | 98 (9.4) | 1 (1.0) | 0.64 (0.08-5.10) | 0.674 | 0.58 (0.07-5.16) | 0.626 |
| **Prior infection status<sup>§</sup>** |  |  |  |  |  |  |  |  |
| No | 312 (91.8) | 7 (2.2) | 953 (91.4) | 9 (0.9) | 1.00 |  | 1.00 |  |
| Yes | 28 (8.2) | 0 (0.0) | 90 (8.6) | 0 (0.0) | 1.00 (1.00-1.00) | -- | 1.00 (1.00-1.00) | -- |
| **Age (years)** |  |  |  |  |  |  |  |  |
| <75 | 308 (90.6) | 5 (1.6) | 1,031 (98.8) | 8 (0.8) | 1.00 |  | 1.00 |  |
| ≥75 | 32 (9.4) | 2 (6.3) | 12 (1.2) | 1 (8.3) | 7.46 (2.05-27.20) | 0.002 | 6.92 (1.50-32.02) | 0.013 |
| **Sex** |  |  |  |  |  |  |  |  |
| Male | 100 (29.4) | 2 (2.0) | 274 (26.3) | 4 (1.5) | 1.00 |  | 1.00 |  |
| Female | 240 (70.6) | 5 (2.1) | 769 (73.7) | 5 (0.7) | 0.61 (0.22-1.70) | 0.348 | 0.61 (0.21-1.74) | 0.357 |
| **Nationality** |  |  |  |  |  |  |  |  |
| Qatari | 116 (34.1) | 2 (1.7) | 483 (46.3) | 3 (0.6) | 1.00 |  | 1.00 |  |
| CMW nationalities<sup>¶</sup> | 74 (21.8) | 2 (2.7) | 159 (15.2) | 0 (0.0) | 1.03 (0.20-5.34) | 0.973 | 1.22 (0.21-6.94) | 0.822 |
| Other nationalities | 150 (44.1) | 3 (2.0) | 401 (38.4) | 6 (1.5) | 1.97 (0.66-5.92) | 0.226 | 2.01 (0.62-6.57) | 0.246 |
| **Epidemic phase<sup>\*\*</sup>** |  |  |  |  |  |  |  |  |
| Delta-dominated incidence | 104 (30.6) | 4 (3.8) | 172 (16.5) | 4 (2.3) | 1.00 |  | 1.00 |  |
| Omicron-dominated incidence | 236 (69.4) | 3 (1.3) | 871 (83.5) | 5 (0.6) | 0.24 (0.09-0.66) | 0.005 | 0.30 (0.10-0.88) | 0.028 |

Abbreviations: AOR, adjusted odds ratio; CI, confidence interval; CMW, craft and manual worker; COVID-19, coronavirus disease 2019; OR, odds ratio.
\*These include immunocompromised individuals (solid organ or hematopoietic stem cell transplant recipients, patients receiving chemotherapy or immunosuppressive treatments, patients with severe immunodeficiency, and HIV patients), unvaccinated individuals, those ≥75 years of age, and pregnant women.
<sup>†</sup>Severity [1], criticality [1], and fatality [2] were defined according to the World Health Organization guidelines.
<sup>‡</sup>Proportion of those who progressed to severe, critical, or fatal COVID-19 among those in predictor category.
<sup>§</sup>Vaccination status and prior infection status were ascertained at time of infection.
<sup>¶</sup>These include Bangladeshis, Indians, Nepalese, Pakistanis, Sri Lankans, and Sudanese due to large proportions of these nationals being craft and manual workers.
<sup>\*\*</sup>Before December 19, 2021 incidence in Qatar was dominated by the Delta variant whereas starting from December 19, 2021 incidence was dominated by the Omicron variant.

